# Pre-diagnostic plasma endogenous steroids and the risk of exfoliation glaucoma

**DOI:** 10.64898/2026.03.22.26348920

**Authors:** Namuunaa Juramt, Zoe Zhiyu Ngo, Danielle E. Haslam, Hannah Hwang, Megan Yu, Oana Zeleznik, Louis R. Pasquale, Janey L. Wiggs, Jessica Lasky-Su, Jae H. Kang

**Author notes:** Co-first authors (Namuunaa Juramt, Zoe Zhiyu Ngo). Corresponding Author: Jae Hee Kang, ScD Brigham and Women’s Hospital 181 Longwood Ave Boston, MA 02115.

## Abstract

**Purpose:** Exfoliation glaucoma (XFG) is the most common secondary glaucoma. Prior studies suggest a higher incidence in women and links to reproductive history, implying estrogen-related pathways. Metabolomic data also indicated inverse associations with steroid-related plasma metabolites, suggesting steroid involvement in XFG pathogenesis.

**Methods:** We conducted a nested case-control study within the Nurses’ Health Study (NHS) (1980–2018), NHSII (1989–2019), and Health Professionals Follow-up Study (1986–2018), with 217 XFG suspect (XFGS)/XFG cases and 217 matched controls (62 men and 372 women). We evaluated 18 endogenous steroids and five steroid classes using conditional logistic regression. Secondary analyses examined effect modifications by age and residential latitude, and heterogeneity by disease severity (XFGS vs. XFG). Metabolite set enrichment analysis (MSEA) was used for class-level associations. Multiple comparisons were addressed using the number of effective tests (NEF) for individual steroids and false discovery rate (FDR) for steroid classes.

**Results:** No individual steroid or steroid class met NEF- or FDR-adjusted significance thresholds, overall or by sex. Nonetheless, across both sexes, MSEA demonstrated a non-significant inverse trend between androgen levels and XFG/XFGS risk (FDR=0.22), with 11-ketotestosterone showing a nominal inverse association (OR=0.54; 95%CI=0.31-0.93; P=0.03). Progestogens showed enrichment scores in the positive trend (FDR=0.31), with a borderline positive association between progesterone and XFG/XFGS (OR=2.21; 95%CI=1.00-4.87; P=0.05).

**Conclusions:** Although we observed no statistically significant associations with steroids after correction for multiple testing, the suggestive patterns for androgens and progestogens support the possibility of steroid-related pathways in XFG etiology and support further evaluation in larger studies.

## Introduction

Exfoliation syndrome (XFS) is a systemic disorder characterized by the production and deposition of exfoliation material (XFM).^1^ EM accumulates systemically but is predominantly diagnosed through a dilated eye exam.^2^ In the eye, EM accumulates in the anterior segment, impeding aqueous humor outflow through the trabecular meshwork thereby contributing to elevated intraocular pressure (IOP).^2^ Elevated IOP, along with XFS, are established risk factors for exfoliation glaucoma (XFG) and XFG suspect (XFGS).^3,4^ Current understanding of XFG etiology remains limited, with only a few recognized risk factors including age, family history, residence at extra-equatorial latitudes, ocular ultraviolet (UV) exposure, and genetic variants in *LOXL1*.^5,6^

To further clarify XFG/XFGS risk pathways, prior metabolomic analyses of 379 metabolites^7^ reported inverse associations with steroids and steroid derivatives, particularly cortisone.^7^ Furthermore, cortisone demonstrated similar non-significant inverse associations with a higher XFS genetic risk score.^8^ Corticosteroids can regulate matrix metalloproteinases involved in EM turnover, which is central to XFS pathobiology.^9^ Elevated cortisone/cortisol levels also influence glucose metabolism and T2D risk,^10^ and T2D has been inversely associated with XFG/XFGS in older adults, potentially via increased glycation of basement membrane components that may inhibit EM aggregation.^11,12^ Furthermore, cortisol/cortisone modulates the synthesis of sex-specific steroid hormones,^13,14^ including testosterone^15^ and estrogen,^16^ offering a biologically plausible link to sex-related differences in XFG/XFGS susceptibility.^17^ Some data suggests that estrogen may interact with LOXL1,^18,19^ and we previously observed a higher incidence in women than in men.^5^ Among women, a reproductive history consistent with greater life-time estrogen exposure,^20^ particularly among those with higher XFS genetic susceptibility, was associated with elevated disease risk.

To further elucidate the role of steroid pathways and risk of XFG/XFGS, we conducted a nested case-control study to evaluate the association between pre-diagnostic plasma endogenous steroid hormones and XFG/XFGS. In contrast to our previous metabolomic studies that evaluated the relative levels of a limited set of steroids, we evaluated the absolute concentrations of 18 steroid hormones measured roughly a decade before diagnosis in our cases and their matched controls, enabling a more direct assessment of steroid pathways in XFG/XFGS etiology.

## Methods

### Study Population and Design

We conducted a 1:1 matched case-control study, nested within the Nurses’ Health Study (NHS), the NHSII, and the Health Professionals Follow-up Study (HPFS; registered on ClinicalTrials.gov (NCT00005182)). The NHS began in 1976 and consists of a cohort of 121,700 U.S. female registered nurses between the age range of 35-55 years. The NHSII was initiated in 1989 and consists of a cohort of 116,429 female registered nurses between the age range of 25-42 years. The HPFS began in 1986 with 51,529 male health professionals who were 40-75 years old. Participants were asked to complete and return a biennial questionnaire updating lifestyle and health information, including any new glaucoma diagnoses. This study was approved by the Institutional Review Board (IRB), Human Research Committees of Brigham & Women’s Hospital, Massachusetts Eye and Ear, and Harvard T.H. Chan School of Public Health. Participants’ return of questionnaires and blood samples was considered implicit consent by the IRB.

We identified 217 incident XFG/XFGS cases through 2016 (average time to diagnosis since blood draw = 11.8 years) who self-reported glaucoma and were confirmed as XFG/XFGS cases with medical record review by a glaucoma expert (LRP). Controls were selected using 1:1 risk-set sampling among those at risk as of the matched cases’ diagnosis date; the 217 controls reported having an eye exam but reported no physician-diagnoses of glaucoma. Controls were matched to cases on age at blood draw, sex, ancestry (Scandinavian, Southern-European Caucasian, other Caucasian, others), residential latitude and longitude at blood draw, time of blood draw, month and year of blood draw, and mean years from blood draw to diagnosis date. Among females, controls were matched on menopausal/hormone therapy use status at blood draw (premenopausal, post-menopausal with hormone therapy use, postmenopausal and no hormone therapy use, missing or unknown) and at diagnosis date. For all matching factors and covariates, data were extracted from the biennial survey questionnaire at the time of blood draw. For participants with missing data, we used the biennial questionnaire immediately before the date of obtaining the blood sample.

### Blood Sample Collection

Blood samples for this study were collected in the following years: 1989-1990 (NHS, n=32,826), 1996-1999 (NHSII, n=29,611), and 1993-1995 (HPFS, n=18,159).^7,21^ Participants were sent a blood collection kit and were asked to mail the samples back. In the NHS and the NHSII, participants were asked to provide two 15-mL sodium heparin tubes containing the blood sample. HPFS participants were asked to provide three 10-mL ethylenediaminetetraacetic acid (EDTA) tubes containing blood samples. These tubes were shipped overnight, on ice, to our laboratory for next-day processing by centrifugation.^22^ Upon arrival, samples were separated into 8 cryotubes consisting of 5 tubes of plasma, 2 tubes of white blood cells, and 1 tube of red blood cells. All samples were stored in a large liquid nitrogen freezer farm at -130 °C.

### Measurement of Steroid Hormone Concentrations in Plasma

Plasma steroid hormones were measured at Precion (Morrisville, North Carolina), using liquid chromatography tandem mass spectrometry (LC-MS/MS). We assayed 18 hormones (3 progestogens [progesterone, 17a-hydroxyprogesterone, pregnenolone sulfate], 4 mineralocorticoids [aldosterone, deoxycorticosterone corticosterone, 18-hydroxycorticosterone], 3 glucocorticoids [cortisol, cortisone, 11-deoxycortisol], 5 androgens [testosterone, androstenedione dehydroepiandrosterone sulfate, 11-ketotestosterone, 11b-hydroxyandrostenedione], and 3 estrogens [estradiol, estrone, estrone sulfate]) (Supplementary Table S1). The steroid hormone panel analysis consists of two consecutive extractions from a single plasma/serum sample (200 µL). The extracts were analyzed with three different LC-MS/MS methods. All steroids were log-transformed for statistical analyses, and all outliers were truncated by sex using the extreme Studentized deviate method.^23^

Aldosterone, estrone, and estradiol were measured using Sciex Exion LC/Sciex 5500+ Triple Quadrupole Mass Spectrometer LC-MS/MS system in ESI *negative* mode using C18 reverse phase chromatography. A second aliquot of the reconstituted extract was analyzed using ESI *positive* mode, C18 reversed phase chromatography, covering all non-conjugated steroids. Lastly, the remaining aqueous phase of the first extraction, consisting of polar steroid sulfates (pregnenolone sulfate, dehydroepiandrosterone sulfate, and estrone sulfate) was subjected to protein precipitation with an organic solvent. This supernatant was then analyzed with Sciex Exion LC/Sciex 5500+ Triple Quadrupole Mass Spectrometer LC-MS/MS system in ESI negative mode using C18 reversed phase chromatography (Supplementary Material).

### Quality Control Measures

For quality control, we conducted three pilot studies (Supplementary Table S2, Supplementary Table S3, and Supplementary Table S4): split pilot, processing method pilot, and within-person stability pilot studies using pooled blood samples from healthy volunteers. Overall, reported mean coefficients of variation (CV) for all 18 included steroids were ≤15%, demonstrating good split-sample reproducibility. Furthermore, all showed a Spearman correlation or interclass correlation (ICC) of ≥0.75, demonstrating good delayed processing reproducibility. Lastly, stable within-person reproducibility over 1-2 years was observed for all 18 steroids with ICC ≥0.40, with the exception of cortisol, which shows high fluctuation and high within-person variability.

### XFG/XFGS Case Ascertainment

In the biennial questionnaire, we asked about new physician-diagnosed glaucoma. From participants responding “yes” to this question, we sought to obtain permission to contact their eye care provider for confirmatory medical information. We then contacted the eye care providers of record to send all visual field (VF) reports and to complete a glaucoma questionnaire, which included questions on the presence of XFM, other secondary causes of elevated IOP, optic nerve features, VF loss, and the status of the filtration apparatus. A glaucoma specialist (LRP) reviewed the records in a systematic manner for confirmation and classification. XFG/XFGS was defined as a participant with an eye containing XFM plus one of the following three glaucomatous signs: (1) a history of IOP ≥22 mm Hg, (2) cup-to-disc ratio ≥0.6 or cup-to-disc ratio ≥0.2 units higher than the fellow eye, or (3) one or more reliable VF tests showing glaucomatous VF loss. Given that the first step in case ascertainment was responding to a question about glaucoma, to minimize bias, participants who were confirmed to only have exfoliation material without any of the 3 glaucomatous signs were censored as of the date of diagnosis.

### Statistical Analysis

We used multivariable-adjusted conditional logistic regression for our primary analyses to compare XFG/XFGS cases vs. risk-set sampled, matched controls in the overall sample and by sex. To evaluate the association between the 18 individual endogenous steroids, the 5 steroid classes (Supplementary Table S1), and the risk of XFG/XFGS, we used two conditional logistic regression models. Due to the non-exact matching between cases and controls, to further account for residual confounding, we included adjustments for the matching factors: mean age at blood draw (years), sex, ancestry (other Caucasian, non-Caucasian, southern Caucasian, Scandinavian Caucasian), time of blood draw, month of blood draw, year of blood draw, fasting status, mean latitude of residence, mean longitude of residence, mean years from blood draw to diagnosis date, current status of menopausal hormone therapy use as of blood draw (among females), current status of menopausal hormone therapy use as of diagnosis date (among females). Model 2 incorporated Model 1 covariates with additional adjustment for mean pack-years of smoking (pack years), body mass index (kg/m2), mean physical activity (metabolic equivalent of task [MET]-hours per week), sunlight exposure during summer in youth (≤5hrs, 6-10hrs, ≥11hrs), nonmelanoma skin cancer as of the date of the blood draw (yes/no), mean population density based on residential census tract (number/km^2^), mean folate intake (mg/day), mean caffeine intake (mg/day), mean alcohol intake (g/day), mean caloric intake (kcal/day), myocardial infarction (yes/no), diabetes (yes/no), self-reported high serum cholesterol (yes/no), mean sleep duration (hours), and oral steroids use (yes/no).

Using conditional logistic regression models, we estimated the risk of XFG/XFGS per one log transformed unit increase in hormone metabolite level. A metabolite set enrichment analysis (MSEA) was used to characterize the biochemical processes contributing to XFG/XFGS among hormones of similar molecular structure or biological characteristics (5 steroid hormone classes). We used the number of effective test^24^ (NEF) approach to adjust for multiple comparisons, as it also accounted for correlations between steroid hormones; an NEF of < 0.05 was considered statistically significant whereas a NEF < 0.2 was considered worthy of further investigation. False discovery rate^25^ (FDR) was used for evaluating steroid classes; an FDR < 0.05 was considered statistically significant.

To evaluate potential effect modification, we investigated several known risk factors of XFG/XFGS such as age (≤59 vs >59 years) and residential latitude (<41°N vs. ≥41°N). Furthermore, we assessed whether associations differed by subtypes defined by disease severity (cases with VF loss (XFG) vs. cases without VF loss (XFGS)).

## Results

Among the 217 cases, the mean age at blood draw was 58.6 years (standard deviation (SD) 6.9 years), mean years from blood draw to diagnosis date was 11.8 (SD 6.4) years, and mean latitude of residence was 39.6°N (SD 4.1) (Table 1). Furthermore, 14.3% of cases were male, 6.9% were Scandinavian Caucasian, 24.4% had a family history of glaucoma, and 86.6% and 91.4% of women were post-menopausal as of the blood draw and diagnosis date, respectively. As of the blood draw, 33.9% of women were users of postmenopausal hormones users, and 29.6% of women were users of menopausal hormones users as of the diagnosis date. Also, compared to female participants, male participants were, on average, older at the time of blood draw (62.0 vs. 57.7 years). As expected, the distribution of most matching factors was similar in cases and controls, but cases had a slightly greater family history of glaucoma (24.4% vs. 18.9%). Also, controls were generally less healthy (higher prevalence of hypertension, diabetes, hypercholesteremia and lower levels of physical activity). Among the 217 cases of XFG/XFGS, the highest untreated IOP was 29.6 mmHg (SD 12.6), and the mean cup to disc ratio was 0.56 (SD 0.20). Additionally, 27.2% had IOP ≥22 mmHg only, 26.7% had optic disc cupping only (with or without IOP elevation), and 46.1% had glaucomatous VF loss.

**Table 1.**
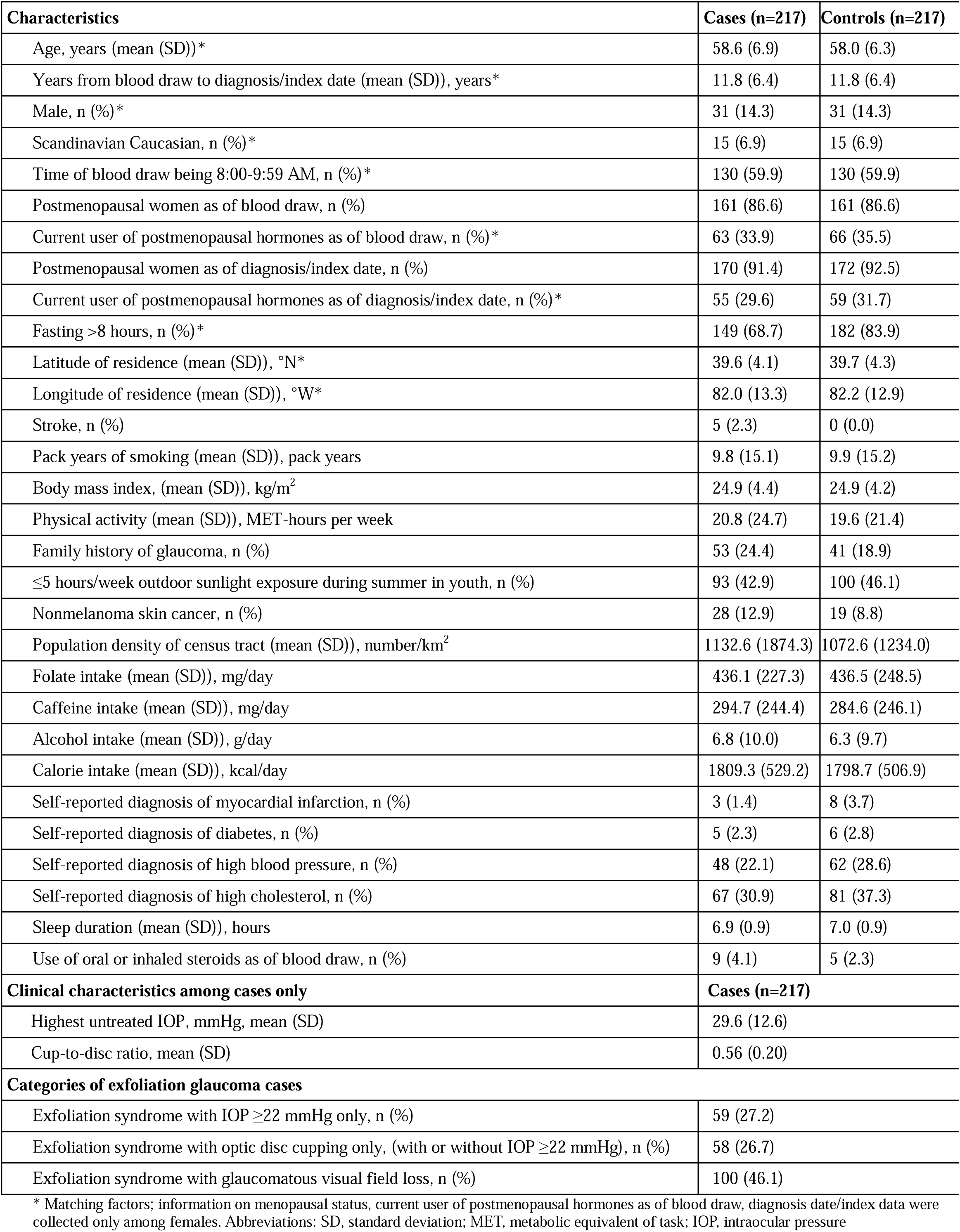
Characteristics of Exfoliation Glaucoma cases and their matched controls.

Table 2 presents the geometric mean of steroids by sex. As expected from prior literature^26^, of the 18 steroids, the distribution of 9 steroids were significantly different by sex (p<0.001); in particular, males had markedly higher testosterone, androstenedione, dehydroepiandrosterone sulfate, estradiol, 17a-hydroxyprogesterone, and pregnenolones sulfate levels (Table 2).

**Table 2.**
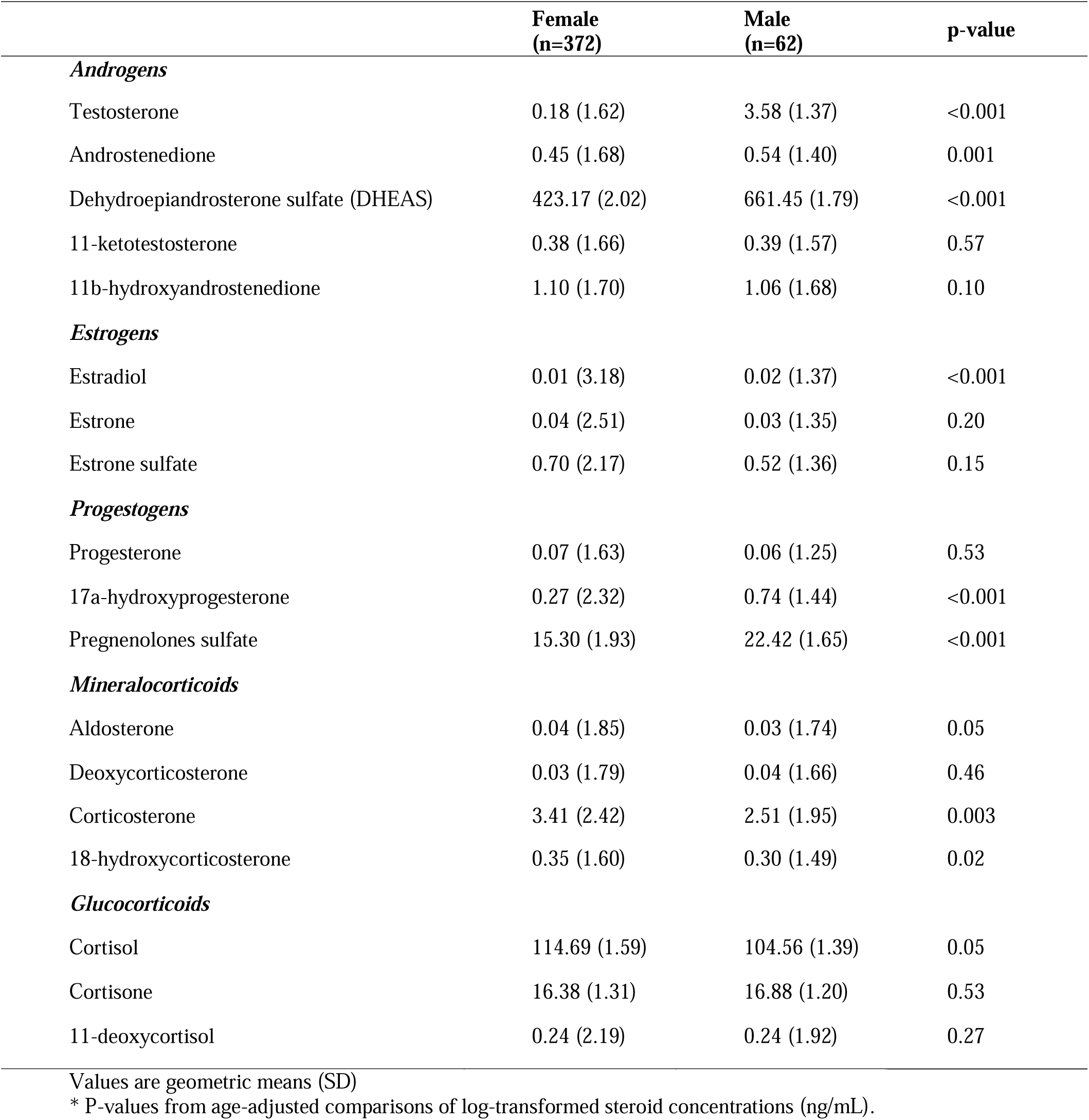
Geometric mean of steroids (ng/mL) by sex*.

|  | **Female**<br>**(n=372)** | **Male**<br>**(n=62)** | **p-value** |
| --- | --- | --- | --- |
| ***Androgens*** |  |  |  |
| Testosterone | 0.18 (1.62) | 3.58 (1.37) | <0.001 |
| Androstenedione | 0.45 (1.68) | 0.54 (1.40) | 0.001 |
| Dehydroepiandrosterone sulfate (DHEAS) | 423.17 (2.02) | 661.45 (1.79) | <0.001 |
| 11-ketotestosterone | 0.38 (1.66) | 0.39 (1.57) | 0.57 |
| 11b-hydroxyandrostenedione | 1.10 (1.70) | 1.06 (1.68) | 0.10 |
| ***Estrogens*** |  |  |  |
| Estradiol | 0.01 (3.18) | 0.02 (1.37) | <0.001 |
| Estrone | 0.04 (2.51) | 0.03 (1.35) | 0.20 |
| Estrone sulfate | 0.70 (2.17) | 0.52 (1.36) | 0.15 |
| ***Progestogens*** |  |  |  |
| Progesterone | 0.07 (1.63) | 0.06 (1.25) | 0.53 |
| 17a-hydroxyprogesterone | 0.27 (2.32) | 0.74 (1.44) | <0.001 |
| Pregnenolones sulfate | 15.30 (1.93) | 22.42 (1.65) | <0.001 |
| ***Mineralocorticoids*** |  |  |  |
| Aldosterone | 0.04 (1.85) | 0.03 (1.74) | 0.05 |
| Deoxycorticosterone | 0.03 (1.79) | 0.04 (1.66) | 0.46 |
| Corticosterone | 3.41 (2.42) | 2.51 (1.95) | 0.003 |
| 18-hydroxycorticosterone | 0.35 (1.60) | 0.30 (1.49) | 0.02 |
| ***Glucocorticoids*** |  |  |  |
| Cortisol | 114.69 (1.59) | 104.56 (1.39) | 0.05 |
| Cortisone | 16.38 (1.31) | 16.88 (1.20) | 0.53 |
| 11-deoxycortisol | 0.24 (2.19) | 0.24 (1.92) | 0.27 |

Values are geometric means (SD)
\* P-values from age-adjusted comparisons of log-transformed steroid concentrations (ng/mL).

### Relationship Between Individual Steroid Hormones and XFG/XFGS

Figure 1 and Supplementary Table S5 presents analyses in the overall sample and in the sex-specific subgroups, respectively. We used two conditional logistic regression models (Model 1 and Model 2); both models yielded comparable results when evaluating the association between 18 individual endogenous steroid hormones (per one log-transformed unit increase in hormone level) in the overall sample. We did not observe any steroids that were significant after NEF multiple correction in the overall sample or in sex-specific strata. Four steroids showed suggestive associations (p≤0.07) in Model 2: 11-ketotestosterone (OR = 0.54 (95% CI: 0.31, 0.93)), testosterone (OR = 0.59 (95% CI: 0.34, 1.04); progesterone (OR = 2.21 (95% CI: 1.00, 4.87)) and cortisone (OR = 0.36 (95% CI: 0.13, 1.03)).

**Figure 1.**
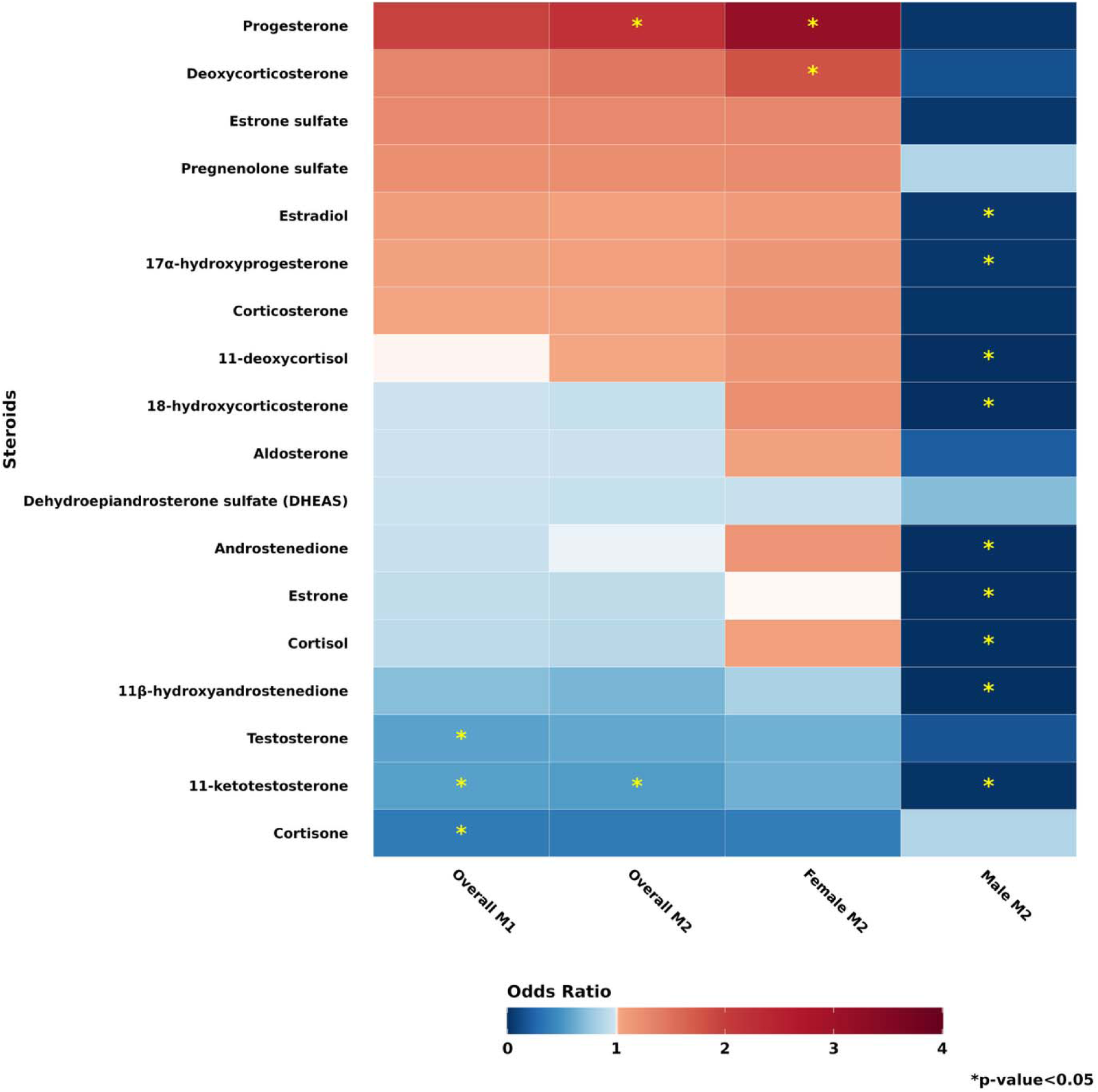
Individual endogenous steroids (n=18) across conditional logistic regression models of exfoliation glaucoma among the overall stud population (217 cases and 217 controls), females (186 cases and 186 controls), and males (31 cases and 31 controls). Model 1 (M1) adjusted for matching factors: mean age at blood draw (years), sex, ancestry (Southern European, Scandinavian, Other Caucasian and non-Caucasian), time of blood draw, year of blood draw, month of blood draw, mean latitude of residence, mean longitude of residence, mean years from blood draw to diagnosis/index date, current status of menopausal hormone therapy use as of blood draw (among females), current status of menopausal hormone therapy use as of diagnosis/index date (among females). Model 2 (M2) adjusted for model 1 and additionally adjusted for family history of glaucoma, mean pack-years of smoking (pack-years), body mass index (kg/m^2^), mean physical activity (metabolic equivalents of task [MET]-hours per week), sunlight exposure during summer in youth (≤5hrs, 6-10hrs, ≥11hrs), nonmelanoma skin cancer as of blood draw (yes/no), mean population density of census tract (number/km^2^), mean folate intake (mg/day), mea caffeine intake (mg/day), mean alcohol intake (g/day), mean caloric intake (kcal/day), comorbidities (0, 1, 2+; comorbidities including myocardial infarction, diabetes, high cholesterol, stroke), mean sleep duration (hours), oral steroids use (yes/no). Class assignment of steroids: Progestogens (progesterone, 17α-hydroxyprogesterone, pregnenolone sulfate); Mineralocorticoids (aldosterone, deoxycorticosterone, corticosterone, 18-hydroxycorticosterone); Glucocorticoids (cortisol, cortisone, 11-deoxycortisol); Androgens (testosterone, androstenedione, dehydroepiandrosterone sulfate, 11-ketotestosterone, 11β-hydroxyandrostenedione); Estrogens (estradiol, estrone, estrone sulfate). P-interaction for sex and steroids ≤0.05 (Model 2): androstenedione (p=0.03), progesterone (p=0.02), 17a-hydroxyprogesterone (p=0.05), 18-hydroxycorticosterone (p=0.03), corticosterone (p=0.03), deoxycorticosterone (p=0.03). * p-value < 0.05.

Among females, Model 2 results showed that progesterone and deoxycorticosterone were significantly positively associated with XFG/XFGS risk. Among male participants, 9 endogenous steroids all showed nominally significant inverse associations with disease risk: 11-ketotestosterone, androstenedione, 11β-hydroxyandrostenedione, 18-hydroxycorticosterone, 11-deoxycortisol, cortisol, estradiol, estrone and 17α-hydroxyprogesterone. Our analysis demonstrated nominally significant interactions by sex in Model 2 for androstenedione (P =0.03), progesterone (P=0.02), 17α-hydroxyprogesterone (P =0.05), 18-hydroxycorticosterone (P=0.03), corticosterone (P=0.03), deoxycorticosterone (P =0.03).

### Relationship Between Steroid Hormone Classes and XFG/XFGS

Using MSEA analysis, both models demonstrated largely consistent patterns of association with XFG/XFGS (Figure 2, Supplementary Table S6). No steroid hormone class met the pre-specified FDR threshold for significance in either model. Nonetheless, progestogen, mineralocorticoid and estrogen classes showed enrichment scores in the positive trend in the overall model, whereas androgens and glucocorticoids demonstrated enrichment scores in the inverse direction (FDR > 0.2). We observed no significant interactions by sex for the MSEA analyses (Figure 2, Supplementary Table S6).

**Figure 2.**
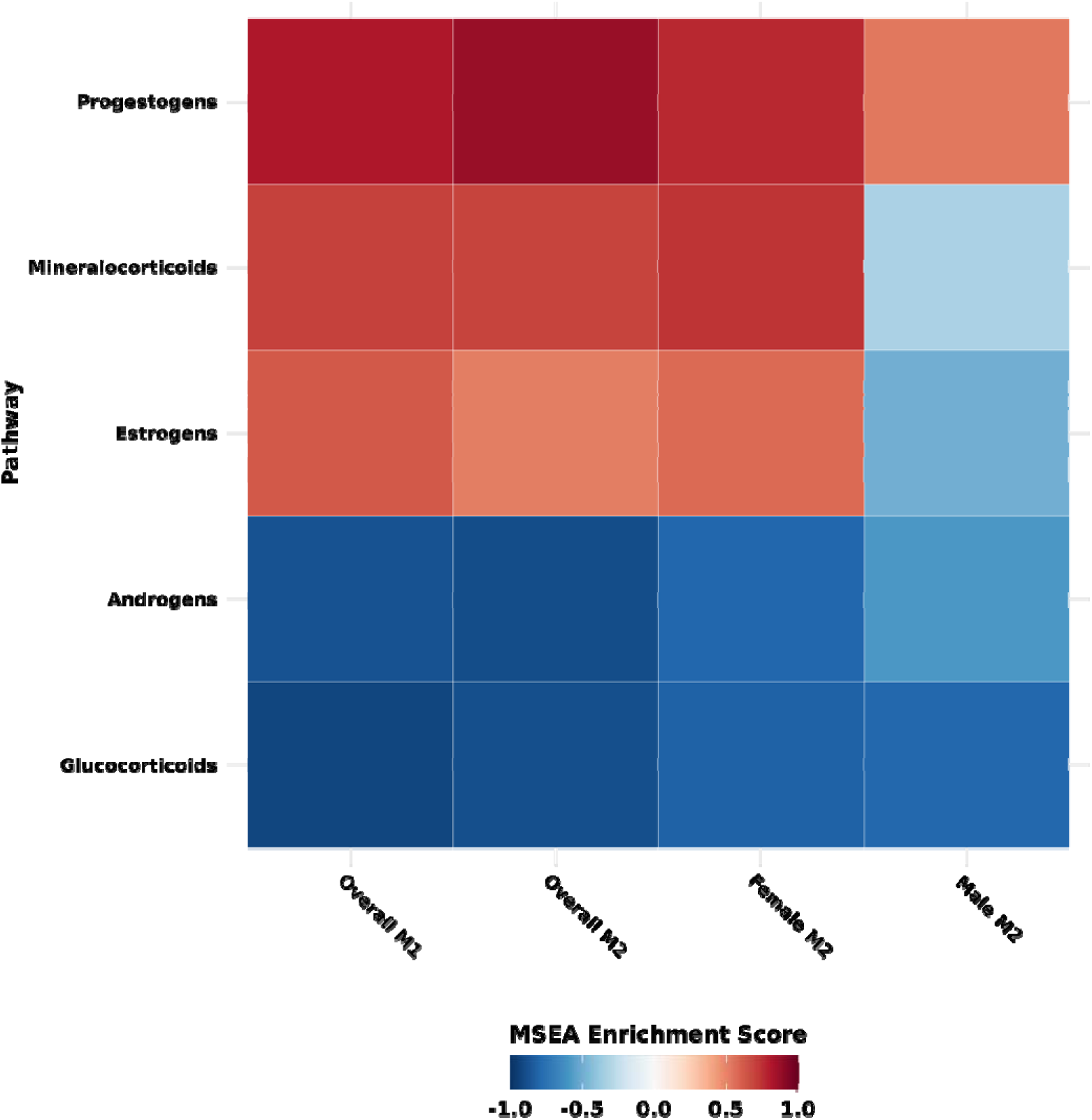
Endogenous steroid classes (n=5) evaluated across conditional logistic regression models of exfoliation glaucoma among the overall study population (217 cases and 217 controls), females (186 cases and 186 controls), and males (31 cases and 31 controls). Model 1 (M1) adjusted for matching factors: mean age at blood draw (years), sex, ancestry (Southern European, Scandinavian, Other Caucasian and non-Caucasian), time of blood draw, year of blood draw, month of blood draw, mean latitude of residence, mean longitude of residence, mean years from blood draw to diagnosis/index date, current status of menopausal hormone therapy use as of blood draw (among females), current status of menopausal hormone therapy use as of diagnosis/index date (among females). Model 2 (M2) adjusted for model 1 and additionally adjusted for family history of glaucoma, mean pack-years of smoking (pack-years), body mass index (kg/m^2^), mean physical activity (metabolic equivalents of task [MET]-hours per week), sunlight exposure during summer in youth (≤5hrs, 6-10hrs, ≥11hrs), nonmelanoma skin cancer as of blood draw (yes/no), mean population density of census tract (number/km^2^), mean folate intake (mg/day), mean caffeine intake (mg/day), mean alcohol intake (g/day), mean caloric intake (kcal/day), comorbidities (0, 1, 2+; comorbidities including myocardial infarction, diabetes, high cholesterol, stroke), mean sleep duration (hours), oral steroids use (yes/no). Class assignment of steroids: Progestogens (progesterone, 17α-hydroxyprogesterone, pregnenolone sulfate); Mineralocorticoids (aldosterone, deoxycorticosterone, corticosterone, 18-hydroxycorticosterone); Glucocorticoids (cortisol, cortisone, 11-deoxycortisol); Androgens (testosterone, androstenedione, dehydroepiandrosterone sulfate, 11-ketotestosterone, 11β-hydroxyandrostenedione); Estrogens (estradiol, estrone, estrone sulfate). MSEA analyses to test for interaction between steroids by sex showed that no class was NEF-significant or NEF<0.2.

### Secondary Analyses

We observed no significant interaction by age or residential latitude across all 18 endogenous steroids (Supplementary Figure S1, Supplementary Figure S2). Analyses evaluating heterogeneity by disease severity (Figure 3) revealed no major differences in the direction of associations for most steroid classes. However, the adverse association with progestogens were FDR-significant for XFG (with visual field loss (VFL)) but not XFGS (without VFL), and a suggestive inverse trend (FDR<0.2) between androgen class and XFG (with VFL), likely driven by 11-ketototestosterone (Supplementary Figure S3).

**Figure 3.**
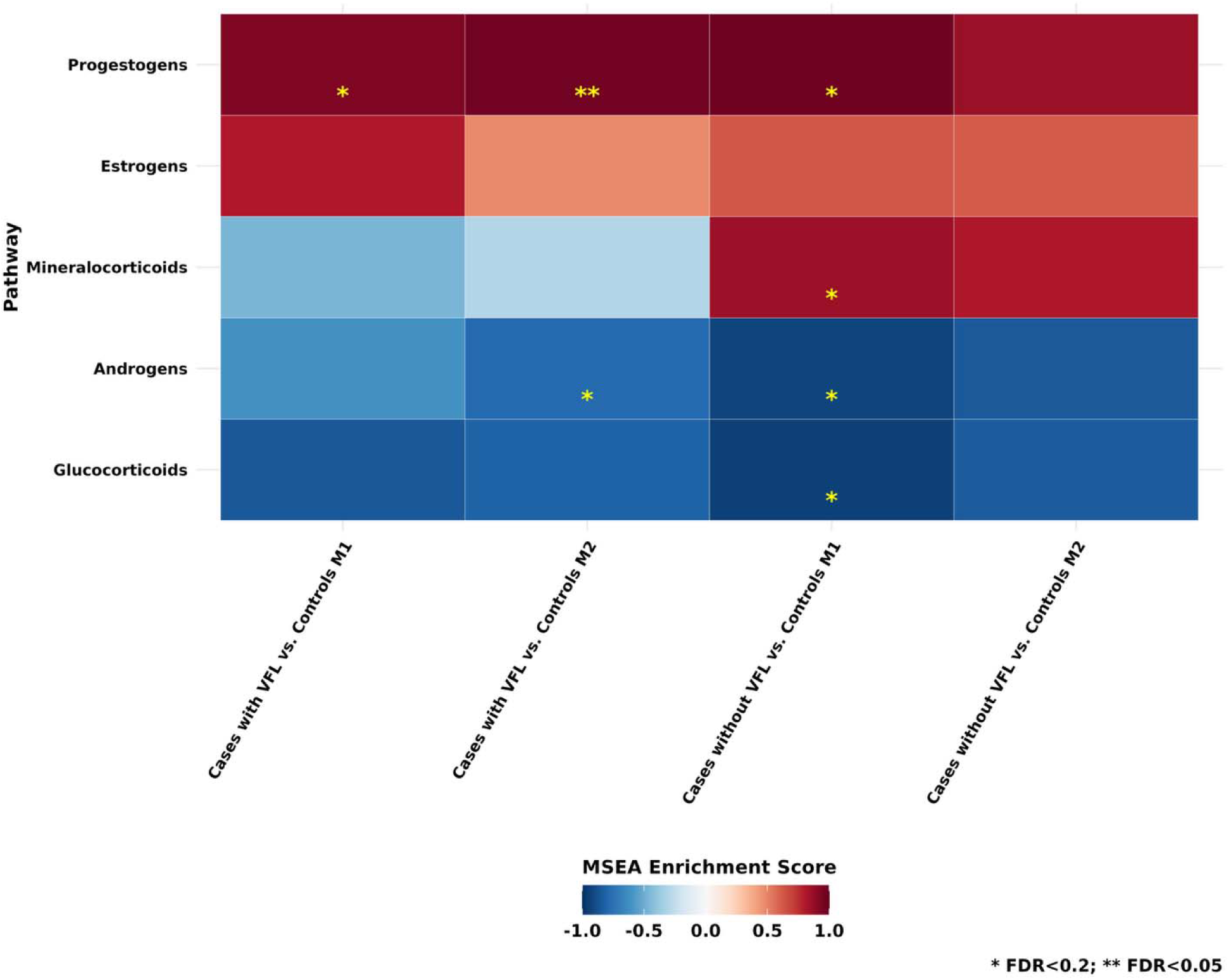
Subtype MSEA analyses by severity of disease: XFG cases with visual field loss (VFL; n=100) vs. controls (n=217); XFGS cases without VFL (n=117) vs. controls (n=217). Model 1 (M1) adjusted for matching factors: mean age at blood draw (years), sex, ancestry (Southern European, Scandinavian, Other Caucasian and non-Caucasian), time of blood draw, year of blood draw, month of blood draw, mean latitude of residence, mean longitude of residence, mean years from blood draw to diagnosis/index date, current status of menopausal hormone therapy use as of blood draw (among females), current status of menopausal hormone therapy use as of diagnosis/index date (among females). Model 2 (M2) adjusted for model 1 and additionally adjusted for family history of glaucoma, mean pack-years of smoking (pack-years), body mass index (kg/m^2^), mean physical activity (metabolic equivalents of task [MET]-hours per week), sunlight exposure during summer in youth (≤5hrs, 6-10hrs, ≥11hrs), nonmelanoma skin cancer as of blood draw (yes/no), mean population density of census tract (number/km^2^), mean folate intake (mg/day), mean caffeine intake (mg/day), mean alcohol intake (g/day), mean caloric intake (kcal/day), comorbidities (0, 1, 2+; comorbidities including myocardial infarction, diabetes, high cholesterol, stroke), mean sleep duration (hours), oral steroids use (yes/no). Class assignment of steroids: Progestogens (progesterone, 17α-hydroxyprogesterone, pregnenolone sulfate); Mineralocorticoids (aldosterone, deoxycorticosterone, corticosterone, 18-hydroxycorticosterone); Glucocorticoids (cortisol, cortisone, 11-deoxycortisol); Androgens (testosterone, androstenedione, dehydroepiandrosterone sulfate, 11-ketotestosterone, 11β-hydroxyandrostenedione); Estrogens (estradiol, estrone, estrone sulfate). Analyses by disease severity (XFG (with VL) versus XFG suspect (without VL) showed no steroids with significant p-heterogeneity.

## Discussion

In this 1:1 matched case-control study nested within the NHS, NHSII, and HPFS evaluating 18 plasma endogenous steroid hormones assessed 11.8 years before diagnosis, we observed no statistically significant associations with the risk of XFG/XFGS overall or by sex after correction for multiple testing. Androgen hormone class (particularly 11-ketotestosterone) showed trends of inverse associations with XFG/XFGS, while progestogens showed trends of adverse associations (FDR < 0.2) that warrant further study in a larger prospective study.

### Androgens

Several androgen-related steroids showed suggestive protective associations with XFG/XFGS risk in males and in the overall cohort. Testosterone, an unoxidized form of 11-ketotestosterone and a downstream product of androstenedione, has been studied in relation to primary open-angle glaucoma, although findings remain mixed.^27,28^ Prior work reported that testosterone can downregulate TGF-β1 signaling in mouse satellite cells,^29^ and TGF-β1 regulates lysyl oxidase (LOX) in eye trabecular meshwork cells as well as various fibrotic disease processes.^30,31^ Lysyl oxidase like-1 (LOXL1) overexpression is observed in the early stages of XFG/XFGS, ^32^ and elevated TGF-β1 has been found in the aqueous humor of XFG/XFGS patients.^33^ Together, these data support a potential model in which higher testosterone may reduce XFG/XFGS risk via downregulating TGF-β1-mediated pathways.

Another plausible mechanism involves inflammation; low testosterone has been associated with an increased production of pro-inflammatory cytokines,^34^ whereas testosterone supplementation has been linked to a reduction in systemic inflammatory markers.^35,36^ Thus, reduced androgen levels may have a potential role in increasing inflammation relevant to XFG disease processes.

### Cortisone

A previous study identified a strong inverse association between cortisone and XFG/XFG, ^7^and another study found similar non-significant inverse association between cortisone and LOXL1-associated XFS genetic risk score.^8^ Importantly, neither study evaluated absolute hormone concentrations, and in the present analysis, we were able to address this limitation by using directly measured absolute hormone levels. Our study observed an inverse association between cortisone and XFG/XFGS risk across all cohorts (male, female, and overall), although results did not reach nominal significance.

Although evidence has been mixed, elevated cortisol levels, the active form of cortisone, has been associated with increased risk of diabetes.^37,38^ A 2024 systematic review and meta-analysis reported that diabetes mellitus was inversely associated with XFG/XFGS in older adults (mean age ≥65 years).^39^ It was hypothesized that chronic hyperglycemia may increase glycation of structural proteins such as fibrinogen and collagen,^12^ potentially disrupting XFM formation and thereby lowering XFG/XFGS risk.

### Progestogens and estrogens

Because our cohort consisted mostly of postmenopausal women whose estrogen and progesterone levels were substantially reduced, the relatively low progestogen and estrogen levels may not have reflected lifetime exposure, potentially contributing to the modest associations observed with these two classes. Nevertheless, we observed that progestogens, particularly progesterone, were associated with an increased risk of XFG/XFGS. Our group previously observed that both earlier age at menarche and natural (versus surgical) menopause, both linked to higher life-time estrogen and progesterone exposure,^40^ were associated with an increased risk of XFS/XFGS.^20^ Progesterone has been found to enhance TGF-β1 expression in epithelial cells,^41^ a key regulator in the disease process, suggestive of a potential mechanistic link. We also observed that higher estrogen was associated with non-significantly higher risk of XFG/XFGS. Prior mice studies have found that 17-β estradiol significantly upregulated LOXL1 and TGF-β1 in mouse urogenital tissues and human Ishikawa cells,^42^ Estrogen treatment significantly upregulated lysyl oxidase expression 12-fold in guinea pigs, whereas low levels of estrogen reduced lysyl oxidase expression toward baseline in non-estrogenized guinea pigs.^43^ These findings collectively suggest that both estrogen and progesterone may influence pathways relevant to XFM formation.

### Strengths and Limitations

This study has several strengths. First, our case definition demonstrates high specificity. This is validated given our prior work highlighting reproducible strong age-related increase in incident XFG, latitude effect on disease risk, and the association between XFS genetic risk score and XFS.^5,8^ Second, participants in these studies are followed biennially, resulting in a comprehensive collection of covariates to minimize the potential for confounding. In particular, we were able to adjust for female reproductive health factors such as the use of post-menopausal hormone therapy. This investigation also builds on our prior work examining pre-diagnostic plasma metabolomics and the risk of XFG/XFGS.^7^ A key limitation of our earlier study was the lack of absolute concentration data for certain metabolites, including cortisone. In the present study, we were able to address this gap, improving the precision of our estimated associations. Finally, blood samples were analyzed using liquid chromatography-tandem mass spectrometry (LC-MS/MS), a method known for its high sensitivity, specificity, and cost-effectiveness.^44^ Furthermore, the steroid measurements produced expected sex-specific distributions in both male and female participants, supporting the rigor and validity of the assay platform.

This study has some limitations. Our cohort is composed primarily of highly educated White health professionals, which may limit the generalizability of our findings to other ancestral groups with different occupational profiles. Several factors may have biased our findings towards the null, in addition to the limited statistical power to detect modest associations such that our results did not meet the stringent criteria required for multiple comparisons. The inherently low sex hormone levels in this middle-aged population may have further constrained our ability to identify robust associations between androgens, estrogens and XFG/XFGS. Minor non-differential misclassification is also possible, as we could not definitively exclude XFG/XFGS in all participants who did not report glaucoma. However, given the low population prevalence of XFG/XFGS in the US population [citation], any under-ascertainment would have affected only a small portion of participants and was unlikely to introduce systematic error that would meaningfully bias our findings. Blood samples were stored for an extended period before analysis, raising the possibility of biomarker degradation. Although cases and controls were matched by the time, date, and year of blood collection, degradation of steroid metabolite could still have biased associations toward the null. Importantly, any degradation would have occurred equally in both groups. Moreover, 17 of 18 biomarkers demonstrated acceptable within person stability in our pilot study supporting temporal stability over 1-2 years (Supplementary Table S4). Prior studies using stored NHS samples have successfully identified associations between endogenous sex hormones and breast cancer, further supporting the validity of these samples.^45^ Additionally, although interlaboratory variability may affect CV estimates for LC-MS/MS, previous studies using NHS and HPFS^46,47^ biospecimens have successfully replicated findings^48,49^ across multiple cohorts and health outcomes, supporting the reliability of the samples for research.

In conclusion, this work supports our prior finding of an inverse association between cortisone and XFG/XFGS risk, although the association was not statistically significant. Overall, no steroid hormone associations remained significant after correction for multiple comparisons, but suggestive trends for androgens and progestogens with XFG/XFGS highlight areas for future investigation in larger studies.

## Data Availability

All data produced in the present study are available upon reasonable request to the authors

## Acknowledgements

This work was supported by the National Institutes of Health [UM1 CA186107, U01 CA167552, R01 EY09611, R01 EY020928 (J.L.W., J.H.K), R01 EY015473 (L.R.P.)]. Dr. Pasquale is also supported by The Glaucoma Foundation (NYC) and the Barry Family Center for the Ophthalmic Artificial Intelligence. The funders had no role in the design or conduct of the study; collection, management, analysis, or interpretation of the data; preparation, review or approval of the manuscript; or the decision to submit the manuscript for publication. This study was accepted as an ARVO (Association for Research in Vision and Ophthalmology) conference abstract in 2025. Financial Disclosures: All authors have declared no conflicts of interest. Unrelated to this work: Dr. Wiggs is a consultant for Allergan, Editas, Maze, Regenxbio, and Avellino. Megan Yu is a contractor for Epidemiologic Research & Methods, LLC.

## Commercial Relationships Disclosure

All authors have declared no commercial relationships.

