## Supplementary materials for "Pre-diagnostic plasma endogenous steroids and the risk of exfoliation glaucoma"

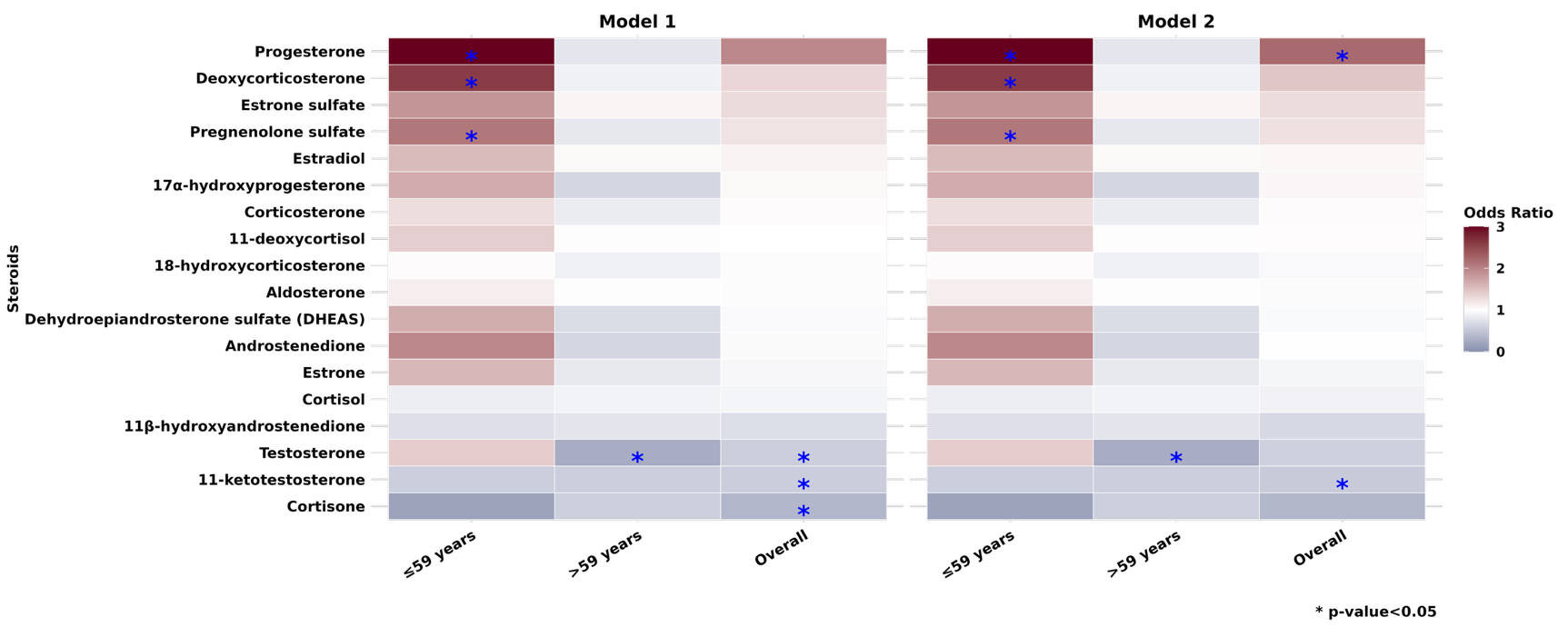

**Supplementary Figure S1.** Secondary analysis for individual endogenous steroids (n=18) by age (≤ 59 (n=215) vs. > 59 years (n=219))

Model 1 (M1) adjusted for matching factors: mean age at blood draw (years), sex, ancestry (Southern European, Scandinavian, Other Caucasian and non-Caucasian), time of blood draw, year of blood draw, month of blood draw, mean latitude of residence, mean longitude of residence, mean years from blood draw to diagnosis/index date, current status of menopausal hormone therapy use as of blood draw (among females), current status of menopausal hormone therapy use as of diagnosis/index date (among females). Model 2 (M2) adjusted for model 1 and additionally adjusted for family history of glaucoma, mean pack-years of smoking (pack-years), body mass index (kg/m^2^), mean physical activity (metabolic equivalents of task [MET]-hours per week), sunlight exposure during summer in youth (≤5hrs, 6-10hrs, ≥11hrs), nonmelanoma skin cancer as of blood draw (yes/no), mean population density of census tract (number/km^2^), mean folate intake (mg/day), mean caffeine intake (mg/day), mean alcohol intake (g/day), mean caloric intake (kcal/day), comorbidities (0, 1, 2+; comorbidities including myocardial infarction, diabetes, high cholesterol, stroke), mean sleep duration (hours), oral steroids use (yes/no). Class assignment of steroids: Progestogens (progesterone, 17α-hydroxyprogesterone, pregnenolone sulfate); Mineralocorticoids (aldosterone, deoxycorticosterone, corticosterone, 18-hydroxycorticosterone); Glucocorticoids (cortisol, cortisone, 11-deoxycortisol); Androgens (testosterone, androstenedione, dehydroepiandrosterone sulfate, 11-ketotestosterone, 11β-hydroxyandrostenedione); Estrogens (estradiol, estrone, estrone sulfate).

No steroid showed a significant interaction (p≤0.05) with age.

* p-value < 0.05.

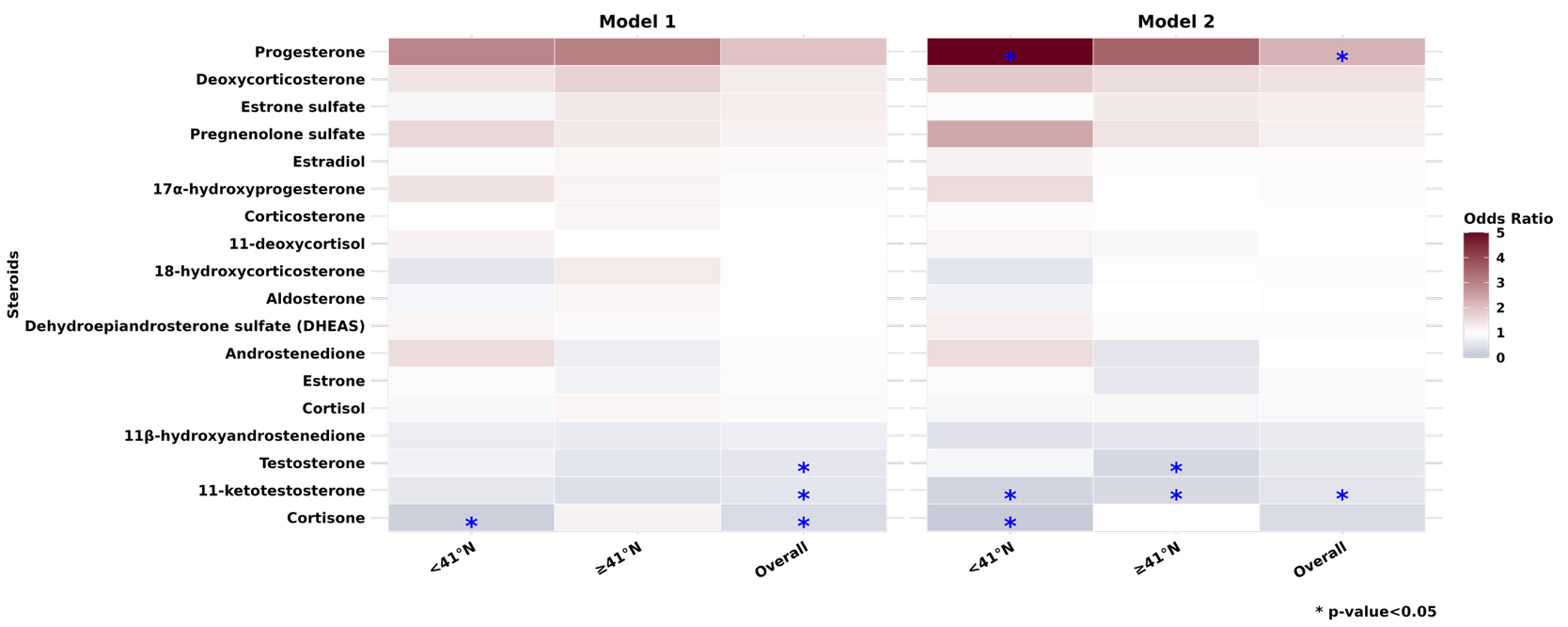

**Supplementary Figure S2.** Secondary analysis for individual endogenous steroids (n=18) by latitude (<41°N (n=196) vs. ≥41°N (n=238))

Model 1 (M1) adjusted for matching factors: mean age at blood draw (years), sex, ancestry (Southern European, Scandinavian, Other Caucasian and non-Caucasian), time of blood draw, year of blood draw, month of blood draw, mean latitude of residence, mean longitude of residence, mean years from blood draw to diagnosis/index date, current status of menopausal hormone therapy use as of blood draw (among females), current status of menopausal hormone therapy use as of diagnosis/index date (among females). Model 2 (M2) adjusted for model 1 and additionally adjusted for family history of glaucoma, mean pack-years of smoking (pack-years), body mass index (kg/m^2^), mean physical activity (metabolic equivalents of task [MET]-hours per week), sunlight exposure during summer in youth (≤5hrs, 6-10hrs, ≥11hrs), nonmelanoma skin cancer as of blood draw (yes/no), mean population density of census tract (number/km^2^), mean folate intake (mg/day), mean caffeine intake (mg/day), mean alcohol intake (g/day), mean caloric intake (kcal/day), comorbidities (0, 1, 2+; comorbidities including myocardial infarction, diabetes, high cholesterol, stroke), mean sleep duration (hours), oral steroids use (yes/no). Class assignment of steroids: Progestogens (progesterone, 17α-hydroxyprogesterone, pregnenolone sulfate); Mineralocorticoids (aldosterone, deoxycorticosterone, corticosterone, 18-hydroxycorticosterone); Glucocorticoids (cortisol, cortisone, 11-deoxycortisol); Androgens (testosterone, androstenedione, dehydroepiandrosterone sulfate, 11-ketotestosterone, 11β-hydroxyandrostenedione); Estrogens (estradiol, estrone, estrone sulfate).

No steroid showed significant interaction (p≤0.05) with latitude.

* p-value < 0.05.

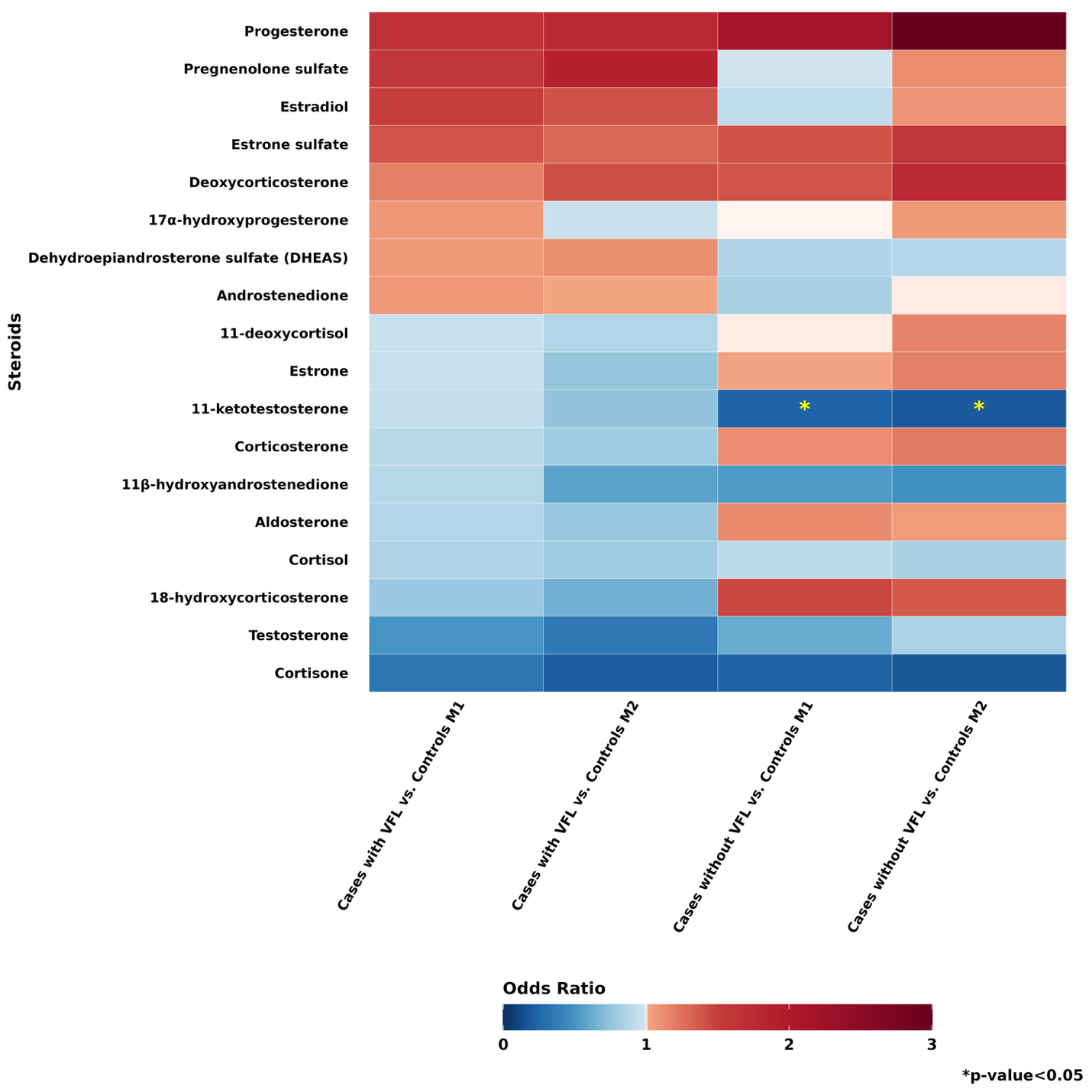

**Supplementary Figure S3.** Subtype analyses by severity of disease: XFG cases with visual field loss (VFL; n=100) vs. controls (n=217); XFGS cases without VFL (n=117) vs. controls (n=217).

Model 1 (M1) adjusted for matching factors: mean age at blood draw (years), sex, ancestry (Southern European, Scandinavian, Other Caucasian and non-Caucasian), time of blood draw, year of blood draw, month of blood draw, mean latitude of residence, mean longitude of residence, mean years from blood draw to diagnosis/index date, current status of menopausal hormone therapy use as of blood draw (among females), current status of menopausal hormone therapy use as of diagnosis/index date (among females). Model 2 (M2) adjusted for model 1 and additionally adjusted for family history of glaucoma, mean pack-years of smoking (pack-years), body mass index (kg/m^2^), mean physical activity (metabolic equivalents of task [MET]-hours per week), sunlight exposure during summer in youth (≤5hrs, 6-10hrs, ≥11hrs), nonmelanoma skin cancer as of blood draw (yes/no), mean population density of census tract (number/km^2^), mean folate intake (mg/day), mean caffeine intake (mg/day), mean alcohol intake (g/day), mean caloric intake (kcal/day), comorbidities (0, 1, 2+; comorbidities including myocardial infarction, diabetes, high cholesterol, stroke), mean sleep duration (hours), oral steroids use (yes/no). Class assignment of steroids: Progestogens (progesterone, 17α-hydroxyprogesterone, pregnenolone sulfate); Mineralocorticoids (aldosterone, deoxycorticosterone, corticosterone, 18-hydroxycorticosterone); Glucocorticoids (cortisol, cortisone, 11-deoxycortisol); Androgens (testosterone, androstenedione, dehydroepiandrosterone sulfate, 11-ketotestosterone, 11β-hydroxyandrostenedione); Estrogens (estradiol, estrone, estrone sulfate).

P-heterogeneity in model 2 by subtype for 11-ketotestosterone was 0.08.

* p-value < 0.05.

**Supplementary Table S1:** List of all Individual Steroid Hormones and Their Classification

**Steroid Hormone Class and Individual Name**

**Progestogens**

Progesterone

17a-hydroxyprogesterone

Pregnenolone sulfate

**Mineralocorticoids**

Aldosterone

Deoxycorticosterone

Corticosterone

18-hydroxycorticosterone

**Glucocorticoids**

Cortisol

Cortisone

11-deoxycortisol

**Androgens**

Testosterone

Androstenedione

Dehydroepiandrosterone sulfate (DHEAS)

11-ketotestosterone

11b-hydroxyandrostenedione

**Estrogens**

Estradiol

Estrone

Estrone sulfate

**Quality Control Measures**

For evaluating the feasibility of the comprehensive steroid hormone panel analyzed in this study using various LC-MS/MS platforms, we conducted three pilot studies (split, processing method (PM), and within-person stability (WPS)). Prednisone and Prednisolone were not considered in the analysis due to unquantifiable samples, leaving 18 hormone panels available to analysis. Quality control studies (QC), such as Split pilot, was used to assess reproducibility across identical samples and determined all 18 hormone panels had good levels of inter-assay reproducibility. In the processing method (PM) pilot, all hormone panels except 11-ketotestosterone received an interclass correlation (ICC) ≥0.75 or spearman r coefficient all passed, showing passing sample stability for most samples. However, the 11-ketotestosterone panel later passed with spearman correlation coefficient 0-24 hours r=0.97 and 0-48hours r=0.92 indicating good stability. Lastly, WPS QC pilot determined that all hormones in the panel, besides cortisol, passed with ICC or Spearman r ≥0.40. These results indicate that most biomarkers remain stable within an individual over the course of one year. For cortisol, ICC=0.19 and Spearman r=0.25 were observed, but this was expected as cortisol tend to have high within person variability. Results from these pilot studies demonstrated that 18 of 20 steroid hormones were available for analysis.

***Split Pilot Study***

To determine the inter-assay reproducibility, we conducted a split pilot quality control study. In this study, blinded duplicate samples were used to assess the coefficient of variation (CV). Here, 20 assays were measured in 14 duplicate donor samples collected in EDTA and heparin tubes. CVs for each sample were calculated by dividing the standard deviation (SD) by the mean content and multiplying by 100 for each sample. CVs less than or equal to 15 were considered to represent good inter-assay reproducibility and pass the split pilot study. Upon removing values below the limit of detection, all CVs assessed in the split pilot study achieved CV <10%, passing the QC. Note that 11-deoxycortisol and estrogen assays had high max CV % and thus should be interpreted with caution.

**Supplementary Table S2:** Split Pilot Study Results (n=14 non-cohort volunteers)

| **Biomarker** | **QC CV %** | **QC Max CV %** | **Donor CV % for EDTA** | **Donor CV % for Heparin** |
| --- | --- | --- | --- | --- |
| 11-deoxycortisol | 8.70 | 30.52 | 9.10 | 8.30 |
| 11-ketotestosterone | 6.74 | 19.61 | 9.10 | 4.38 |
| 11b-hydroxyandrostenedione | 3.03 | 10.22 | 2.04 | 4.02 |
| 17a-hydroxyprogesterone | 5.14 | 16.64 | 3.57 | 6.72 |
| 18-hydroxycorticosterone | 5.86 | 15.17 | 4.60 | 6.61 |
| Prednisolone | . | . | . | . |
| Prednisone | . | . | . | . |
| Aldosterone | 3.25 | 8.56 | 3.72 | 2.78 |
| Androstenedione | 3.18 | 8.61 | 2.68 | 3.68 |
| Corticosterone | 4.77 | 11.97 | 4.09 | 5.44 |
| Cortisol | 4.12 | 9.92 | 4.74 | 3.51 |
| Cortisone | 3.58 | 7.79 | 3.29 | 3.87 |
| Dehydroepiandrosterone sulfate | 4.07 | 19.31 | 2.82 | 5.31 |
| Deoxycorticosterone | 7.78 | 28.86 | 6.12 | 8.61 |
| Estradiol | 9.72 | 30.89 | 9.60 | 9.83 |
| Estrone | 3.94 | 10.66 | 3.51 | 4.37 |
| Estrone sulfate | 6.46 | 10.39 | 7.44 | 5.73 |
| Pregnenolone sulfate | 1.70 | 7.03 | 2.46 | 0.95 |
| Progesterone | 2.68 | 8.43 | 1.99 | 3.02 |
| Testosterone | 3.82 | 12.10 | 4.69 | 2.62 |

Passing criteria requires 50% sample to have coefﬁcients of variation (CVs) <0.20 above the limit of detection (LOD). Values were not detectable (below LOD) for prednisolone and prednisone so CVs were not evaluated for these steroids.

Abbreviations: QC, quality control; CV, coefficient of variation; LOD, limit of detection

***Processing Method (PM) Pilot Study***

To ensure the reproducibility of hormones examined in this study over processing delays of 24-48 hours, we computed interclass correlation coefficients (ICCs) for the same non-cohort donor samples. These samples were processed at three timepoints: (1) immediately upon receiving sample, (2) chilled as whole blood with a cool pack and processed 24 hours after receiving sample, (3) chilled as whole blood with a cool pack and processed 48 hours after receiving sample. ICCs are defined as the between-person variance divided by the sum of the within and between person variances. To improve normality of the distributions, the samples were natural log transformed for ICC calculations. For the PM pilot study, an ICC≥0.75 or at least one of the spearman r≥0.75 was considered acceptable.

In this pilot, most assays demonstrated good stability after experiencing delays in processing. Values were not detectable (below LOD) for prednisolone and prednisone so ICCs were not evaluated for these steroids. Most ICCs were ≥0.75 with the exception of 11-ketotestosterone. However, when we looked at the Spearman correlation coefficients for the 11-ketotestosterone assay, it passed with 0-24hr r=0.97 and 0-48hr r=0.92. Thus, this pilot demonstrates good sample stability for processing and analysis if delays in processing were to occur.

**Supplementary Table S3**. Processing Methods Pilot Study Results (n=14 non-cohort volunteers)

| **Biomarker** | **ICC (Log-transformed values)** |
| --- | --- |
| 11-deoxycortisol | 0.95 |
| 11-ketotestosterone | 0.69 |
| 11b-hydroxyandrostenedione | 0.96 |
| 17a-hydroxyprogesterone | 0.99 |
| 18-hydroxycorticosterone | 0.98 |
| Prednisolone | . |
| Prednisone | . |
| Aldosterone | 0.92 |
| Androstenedione | 0.98 |
| Corticosterone | 0.97 |
| Cortisol | 0.98 |
| Cortisone | 0.96 |
| Dehydroepiandrosterone sulfate | 0.82 |
| Deoxycorticosterone | 0.80 |
| Estradiol | 0.83 |
| Estrone | 0.91 |
| Estrone sulfate | 0.80 |
| Pregnenolone sulfate | 0.77 |
| Progesterone | 0.998 |
| Testosterone | 0.91 |

Passing criteria requires 50% sample to have CVs <20%, intraclass correlation coefﬁcient (ICC) or spearman correlation coefficient ≥0.75 above the LOD.

Abbreviations: ICC, intraclass correlation coefficient; CV, coefficient of variation; LOD, limit of detection

***Within Person Stability (WPS) Pilot Study***

To assess the reproducibility of hormone measures within an individual over 1 to 2 years, we examined samples from the same person at two timepoints 1 to 2 years apart. This pilot study aims to assess whether one measure represents long-term exposure. To examine the WPS over 1 to 2 years, Spearman r and ICCs were calculated.

In this study, all biomarkers, except cortisol, passed the WPS stability pilot study, where either the ICC or Spearman r was ≥0.40. High spearman r indicated stability of the sample for one individual over the course of 1 to 2 years. Specifically for cortisol, the ICC=0.19 and the Spearman r=0.25. However, this may be expected as cortisol tends to fluctuate over time, contributing to within-person variability.

**Supplementary Table S4.** Within-Person Stability Pilot Study Results (n=20 NHS; n=20 HPFS)

| **Biomarker** | **ICC (raw)** | **Spearman r** |
| --- | --- | --- |
| 11-deoxycortisol (ng/mL) | 0.44 | 0.55 |
| 17a-hydroxyprogesterone (ng/mL) | 0.62 | 0.32 |
| 18-hydroxycorticosterone (ng/mL) | 0.74 | 0.76 |
| Aldosterone (ng/mL) | 0.45 | 0.52 |
| Androstenedione (ng/mL) | 0.51 | 0.55 |
| Corticosterone (ng/mL) | 0.44 | 0.62 |
| Cortisol (ng/mL) | 0.19 | 0.25 |
| Cortisone (ng/mL) | 0.40 | 0.48 |
| Deoxycorticosterone (ng/mL) | 0.66 | 0.68 |
| DHEA sulfate (ng/mL) | 0.74 | 0.89 |
| Estradiol (ng/mL) | 0.81 | 0.62 |
| Estrone (ng/mL) | 0.78 | 0.90 |
| Estrone sulfate (ng/mL) | 0.58 | 0.82 |
| Pregnenolone sulfate (ng/mL) | 0.83 | 0.81 |
| Progesterone (ng/mL) | 0.61 | 0.63 |
| Testosterone (ng/mL) | 0.60 | 0.19 |
| 11-ketotestosterone (ng/mL) | 0.89 | 0.65 |
| 11b-hydroxyandrostenedione (ng/mL) | 0.69 | 0.87 |

Passing criteria requires 50% sample to have either the ICC or Spearman r ≥0.40 above the LOD.

Abbreviations: ICC, intraclass correlation coefficient; LOD, level of detection

**Supplementary Table S5: Individual endogenous steroids across conditional logistic regression models of exfoliation glaucoma among overall study population, female and male**

| Steroids | Model 1 Overall Odds Ratio (95% CI); p value | Model 2 Overall Odds Ratio (95% CI); p value | Model 2 Female-specific Odds Ratio (95% CI); p value | Model 2 Male-specific Odds Ratio (95% CI); p value | ^a^ P_interaction_  _by sex_ |
| --- | --- | --- | --- | --- | --- |
| Progesterone | 1.95 (0.94, 4.05); 0.07 | 2.21 (1.00, 4.87); 0.05 | 3.18 (1.31, 7.71); 0.01 | 0.03 (3.17 ×10⁻⁴, 2.38);  0.11 | 0.02 |
| Deoxycorticosterone | 1.32 (0.90, 1.94); 0.16 | 1.45 (0.94, 2.22); 0.09 | 1.80 (1.11, 2.91); 0.02 | 0.15 (0.02, 1.13);  0.07 | 0.03 |
| Estrone sulfate | 1.28 (0.84, 1.95); 0.25 | 1.28 (0.81, 2.00); 0.29 | 1.29 (0.81, 2.06); 0.28 | 0.04 (6.66 ×10⁻⁴, 1.92);  0.10 | 0.62 |
| Pregnenolone sulfate | 1.22 (0.85, 1.74); 0.28 | 1.24 (0.84, 1.82); 0.27 | 1.26 (0.83, 1.91); 0.27 | 0.87 (0.21, 3.62);  0.84 | 0.76 |
| Estradiol | 1.09 (0.77, 1.55); 0.62 | 1.06 (0.73, 1.54); 0.76 | 1.11 (0.75, 1.66); 0.60 | 0.04 (1.28 ×10⁻³, 0.96);  0.047 | 0.09 |
| 17α-hydroxyprogesterone | 1.05 (0.77, 1.44); 0.74 | 1.07 (0.77, 1.50); 0.67 | 1.16 (0.83, 1.64); 0.39 | 0.04 (1.72 ×10⁻³, 0.82);  0.04 | 0.05 |
| Corticosterone | 1.03 (0.79, 1.34); 0.84 | 1.03 (0.78, 1.37); 0.82 | 1.19 (0.87, 1.62); 0.27 | 0.02 (2.82 ×10⁻⁴, 1.56);  0.08 | 0.03 |
| 11-deoxycortisol | 1.00 (0.75, 1.35); 0.99 | 1.02 (0.74, 1.40); 0.90 | 1.16 (0.82, 1.64); 0.39 | 7.98×10⁻⁶ (8.39×10⁻¹¹, 0.76);  0.04 | 0.12 |
| 18-hydroxycorticosterone | 0.97 (0.60, 1.59); 0.92 | 0.95 (0.57, 1.59); 0.85 | 1.22 (0.71, 2.11); 0.47 | 1.48 ×10⁻³ (4.06 ×10⁻⁶, 0.54);  0.03 | 0.03 |
| Aldosterone | 0.97 (0.67, 1.41); 0.88 | 0.97 (0.65, 1.44); 0.87 | 1.05 (0.69, 1.59); 0.82 | 0.21 (0.01, 3.08);  0.25 | 0.60 |
| Dehydroepiandrosterone  sulfate (DHEAS) | 0.96 (0.68, 1.36); 0.82 | 0.95 (0.66, 1.37); 0.80 | 0.96 (0.64, 1.43); 0.83 | 0.70 (0.20, 2.48);  0.59 | 0.49 |
| Androstenedione | 0.96 (0.61, 1.49); 0.85 | 1.00 (0.62, 1.60); 0.99 | 1.18 (0.71, 1.94); 0.52 | 2.77 ×10⁻⁴ (2.27 ×10⁻⁷, 0.34);  0.02 | 0.03 |
| Cortisol | 0.91 (0.55, 1.53); 0.73 | 0.89 (0.52, 1.54); 0.68 | 1.07 (0.59, 1.94); 0.81 | 3.63 ×10⁻³ (2.07 ×10⁻⁵, 0.63);  0.03 | 0.07 |
| Estrone | 0.93 (0.65, 1.34); 0.70 | 0.92 (0.62, 1.36); 0.68 | 1.00 (0.67, 1.51); 0.99 | 2.34 ×10⁻⁴ (6.80 ×10⁻⁸, 0.80);  0.04 | 0.09 |
| 11β-hydroxyandrostenedione | 0.71 (0.45, 1.12); 0.14 | 0.67 (0.41, 1.09); 0.11 | 0.84 (0.50, 1.43); 0.53 | 3.92 ×10⁻³ (2.28 ×10⁻⁵, 0.67);  0.03 | 0.08 |
| 11-ketotestosterone | 0.56 (0.34, 0.94); 0.03 | 0.54 (0.31, 0.93); 0.03 | 0.64 (0.36, 1.13); 0.12 | 0.02 (5.44 ×10⁻⁴, 0.64);  0.03 | 0.23 |
| Testosterone | 0.57 (0.34, 0.96); 0.03 | 0.59 (0.34, 1.04); 0.07 | 0.64 (0.36, 1.14); 0.13 | 0.17 (2.49 ×10⁻³, 10.99);  0.40 | 0.12 |
| Cortisone | 0.36 (0.14, 0.98); 0.045 | 0.36 (0.13, 1.03); 0.06 | 0.38 (0.13, 1.13); 0.08 | 0.87 (8.69 ×10⁻⁴, 8.64 ×10²);  0.97 | 0.94 |

^a^ P for interaction for sex and steroids

Model 1 (M1) adjusted for matching factors: mean age at blood draw (years), sex, ancestry (Southern European, Scandinavian, Other Caucasian and non-Caucasian), time of blood draw, year of blood draw, month of blood draw, mean latitude of residence, mean longitude of residence, mean years from blood draw to diagnosis/index date, current status of menopausal hormone therapy use as of blood draw (among females), current status of menopausal hormone therapy use as of diagnosis/index date (among females). Model 2 (M2) adjusted for model 1 + family history of glaucoma, mean pack-years of smoking (pack years), body mass index (kg/m^2^), mean physical activity (MET-hours per week), sunlight exposure during summer in youth (≤5hrs, 6-10hrs, ≥11hrs), nonmelanoma skin cancer as of blood draw (yes/no), mean population density of census tract (number/km^2^), mean folate intake (mg/day), mean caffeine intake (mg/day), mean alcohol intake (g/day), mean caloric intake (kcal/day), comorbidities (0, 1, 2+; comorbidities including myocardial infarction, diabetes, high cholesterol, stroke), mean sleep duration (hours), oral steroids use (yes/no). Class assignment of steroids: Progestogens (progesterone, 17α-hydroxyprogesterone, pregnenolone sulfate); Mineralocorticoids (aldosterone, deoxycorticosterone, corticosterone, 18-hydroxycorticosterone); Glucocorticoids (cortisol, cortisone, 11-deoxycortisol); Androgens (testosterone, androstenedione, dehydroepiandrosterone sulfate, 11-ketotestosterone, 11β-hydroxyandrostenedione); Estrogens (estradiol, estrone, estrone sulfate). p-interaction for sex and steroids ≤0.05 (model 2): progesterone (p=0.02), androstenedione (p=0.03), 17a-hydroxyprogesterone (p=0.05), 18-hydroxycorticosterone (p=0.03), deoxycorticosterone (p=0.03). corticosterone (p=0.03).

**Supplementary Table S6: Endogenous steroid classes evaluated across conditional logistic regression models of exfoliation glaucoma among overall study population, female, males**

| Steroid Classes | Model 1 Overall | Model 2 Overall | Model 2 Female-specific | Model 2 Male-specific |
| --- | --- | --- | --- | --- |
| **Progestogens** |  |  |  |  |
| Overall ES | 0.81 | 0.87 | 0.77 | 0.53 |
| Standard log-2 error | 0.12 | 0.15 | 0.10 | 0.11 |
| Uncorrected p value | 0.29 | 0.18 | 0.29 | 0.56 |
| FDR-corrected p value | 0.45 | 0.31 | 0.36 | 0.92 |
| **Mineralocorticoids** |  |  |  |  |
| Overall ES | 0.70 | 0.69 | 0.74 | -0.32 |
| Standard log-2 error | 0.11 | 0.11 | 0.11 | 0.03 |
| Uncorrected p value | 0.36 | 0.34 | 0.26 | 0.97 |
| FDR-corrected p value | 0.45 | 0.42 | 0.36 | 0.97 |
| **Estrogens** |  |  |  |  |
| Overall ES | 0.63 | 0.52 | 0.57 | -0.49 |
| Standard log-2 error | 0.08 | 0.06 | 0.06 | 0.04 |
| Uncorrected p value | 0.61 | 0.76 | 0.65 | 0.74 |
| FDR-corrected p value | 0.61 | 0.76 | 0.65 | 0.92 |
| **Glucocorticoids** |  |  |  |  |
| Overall ES | -0.92 | -0.88 | -0.81 | -0.79 |
| Standard log-2 error | 0.21 | 0.14 | 0.16 | 0.15 |
| Uncorrected p value | 0.08 | 0.16 | 0.16 | 0.11 |
| FDR-corrected p value | 0.21 | 0.31 | 0.36 | 0.56 |
| **Androgens** |  |  |  |  |
| Overall ES | -0.87 | -0.89 | -0.80 | -0.58 |
| Standard log-2 error | 0.20 | 0.28 | 0.25 | 0.07 |
| Uncorrected p value | 0.08 | 0.04 | 0.08 | 0.39 |
| FDR-corrected p value | 0.21 | 0.22 | 0.36 | 0.92 |

Model 1 (M1) adjusted for matching factors: mean age at blood draw (years), sex, ancestry (Southern European, Scandinavian, Other Caucasian and non-Caucasian), time of blood draw, year of blood draw, month of blood draw, mean latitude of residence, mean longitude of residence, mean years from blood draw to diagnosis/index date, current status of menopausal hormone therapy use as of blood draw (among females), current status of menopausal hormone therapy use as of diagnosis/index date (among females). Model 2 (M2) adjusted for model 1 and additionally adjusted for family history of glaucoma, mean pack-years of smoking (pack-years), body mass index (kg/m^2^), mean physical activity (metabolic equivalents of task [MET]-hours per week), sunlight exposure during summer in youth (≤5hrs, 6-10hrs, ≥11hrs), nonmelanoma skin cancer as of blood draw (yes/no), mean population density of census tract (number/km^2^), mean folate intake (mg/day), mean caffeine intake (mg/day), mean alcohol intake (g/day), mean caloric intake (kcal/day), comorbidities (0, 1, 2+; comorbidities including myocardial infarction, diabetes, high cholesterol, stroke), mean sleep duration (hours), oral steroids use (yes/no). Class assignment of steroids: Progestogens (progesterone, 17α-hydroxyprogesterone, pregnenolone sulfate); Mineralocorticoids (aldosterone, deoxycorticosterone, corticosterone, 18-hydroxycorticosterone); Glucocorticoids (cortisol, cortisone, 11-deoxycortisol); Androgens (testosterone, androstenedione, dehydroepiandrosterone sulfate, 11-ketotestosterone, 11β-hydroxyandrostenedione); Estrogens (estradiol, estrone, estrone sulfate).

Abbreviation: ES, Enrichment Score; FDR, False-discovery rate
